# First detection and report of SARS-CoV-2 Spike protein N501Y mutations in Oklahoma USA

**DOI:** 10.1101/2021.01.26.21250584

**Authors:** Sai Narayanan, Girish Patil, Sunil More, Jeremiah Saliki, Anil Kaul, Akhilesh Ramachandran

## Abstract

We describe the detection of SARS-CoV-2 (VOC)B.1.1.7 lineage in Oklahoma, USA. Various mutations in the S gene and ORF8 with similarity to the genome of B.1.1.7 lineage were detected in 4 of the 6 genomes sequenced and reported here. The sequences have been made available in GISAID. Presence of novel lineages indicate the need for frequent whole genome sequencing to better understand pathogen dynamics in different geographical locations.

---

Since the start of the COVID-19 pandemic in December 2019, several lineages/variants of SARS-CoV-2 have been identified based on whole genome sequencing. As of January 21, 2021 approximately 420,000 whole genome sequences have been deposited in the GISAID database (https://www.gisaid.org/) (1,2). Recently a novel SARS-CoV-2 lineage (B 1.1.7.) was identified in Kent and Greater London in the United Kingdom (3) and is reported to have enhanced transmissibility. Infections by the B.1.1.7 lineage has since been reported from several other countries including the United States (4). We report the identification of B.1.1.7 in Oklahoma, USA.

COVID-19 was first reported in Oklahoma on March 6^th^ 2020 (5). As of January 19^th,^ 2021, a total of 358,374 cases and 3,037 deaths have been registered in the state (6). Six clinical samples, from different counties in the state, collected between January 1^st^ and 8^th^, 2021, were sequenced for this report. RNA was extracted using a commercial kit (MVP, ThermoFisher) and viral presence was detected by real-time PCR using TaqPath COVID-19 kit (Thermofisher, MA, USA) that targets the S gene, N gene and ORF 1ab. The S gene target of all the samples selected for sequencing either failed to amplify or had Ct values that were higher when compared to N gene and ORF1ab. Sequencing was performed using the MinION platform (Oxford Nanopore Technologies, UK) following the ARTIC protocol (7).

Sequenced genomes were assembled using canu (8). Reference assisted consensus assemblies were generated using minimap2 (9) and nanopolish (10), SARS-CoV-2 reference genome Wuhan-Hu-1 (GenBank ID: MN908947.3) was used for reference assembly. Individual genes were annotated using VIGOR (11). Annotated genes and the whole genomes were aligned using MUSCLE aligner in MEGA-X (12). The genome sequences were submitted to GISAID (1) and their GISAID classification and PANGO lineages (13) are shown in table 1. Four of the six samples sequenced carried SNPs similar to those described in the B.1.1.7 lineage (20I/501Y.V1 Variant of Concern (VOC) 202012/01) (4), including N501Y, 69/70 deletion, Y144 deletion, A570D, P681H, T716I, S982A, D1118H mutations. Q27Stop mutation in ORF 8 was detected in 3 out of these 4 samples. N501Y and the previously described D614G mutations (14,15) were detected in all 6 samples (Table 2).

**Table 1:** Genome coverage of SARS-CoV-2 sequenced from different samples. GISAID Accession No., Clade and PANGO (Phylogenetic Assignment of Named Global Outbreak) Lineage from GISAID are shown.

| Genome | Coverage | GISAIID Accession No. | GISAIID Clade | PANGO Lineage |
| --- | --- | --- | --- | --- |
| Oklahoma-ADDL-6 | 36932.1 × | EPI_ISL_812346 | GR | B.1.1.7 |
| Oklahoma-ADDL-7 | 2080.4 × | EPI_ISL_861422 | G | B.1.1.7 |
| Oklahoma-ADDL-8 | 1963.7 × | EPI_ISL_861423 | GR | B.1.1.7 |
| Oklahoma-ADDL-9 | 2192.3 × | EPI_ISL_861424 | G | B.1.1.7 |
| Oklahoma-ADDL-10 | 3992.5 × | EPI_ISL_861425 | GH | B.1 |
| Oklahoma-ADDL-11 | 4399.6 × | EPI_ISL_861426 | GH | B.1 |

**Table 2:** Selected mutations detected in S gene and ORF 8, and their respective amino acid changes when compared to NCBI reference genome (SARS-CoV-2, Wuhan Hu-1, NC 045512.2).

| Mutations observed<br>(Nucleotide position) | Gene | Genomes with mutation |
| --- | --- | --- |
| N501Y (A23063T) | S gene | Oklahoma-ADDL-6, 7,8,9,10,11 |
| A570D (C23271A) | S gene | Oklahoma-ADDL-6,7,9 |
| D614G (A23403G) | S gene | Oklahoma-ADDL-6, 7,8,9,10,11 |
| P681H (C23604A) | S gene | Oklahoma-ADDL-6,7,8,9 |
| T716I (C23709T) | S gene | Oklahoma-ADDL-6,7,8,9 |
| S982A (T24506G) | S gene | Oklahoma-ADDL-6,7,8,9 |
| D1118H (G24914C) | S gene | Oklahoma-ADDL-6 |
| 69/70 (21765 - 21770) DEL | S gene | Oklahoma-ADDL-6,7,8,9 |
| Y144 DEL (21991 - 21993) | S gene | Oklahoma-ADDL-6,7,8,9 |
| Q27Stop (C27972T) | ORF 8 | Oklahoma-ADDL-6,7,8 |

Frequent whole genome sequencing is critical to monitor mutations in the viral genome. The evolution of highly transmissible strains of SARS-CoV-2 and its global spread underlines the need to remain vigilant and continue practicing necessary social measures to prevent further disease spread.

## Data Availability

Sequence data has been made available in GISAID (www.gisaid.org). The accession numbers for the sequences have been provided in the manuscript.

## Ethics Statement

This study was approved by the Institutional Review Board (Application number: IRB-20-357) at Oklahoma State University, Stillwater OK 74078, United States.

## Acknowledgment

We thank Dr. Yiwei Wang and Dr. Jeremy Kaplan for their help in sample collection. Study was partially funded by FDA-CVM-VetLIRN (Grant no.5U18FD006671-02).

